# Exploring Temperature Effects on Large Language Models Across Various Clinical Tasks

**DOI:** 10.1101/2024.07.22.24310824

**Authors:** Dhavalkumar Patel, Prem Timsina, Ganesh Raut, Robert Freeman, Matthew A levin, Girish N Nadkarni, Benjamin S Glicksberg, Eyal Klang

## Abstract

Large Language Models (LLMs) are becoming integral to healthcare analytics. However, the influence of the temperature hyperparameter, which controls output randomness, remains poorly understood in clinical tasks. This study evaluates the effects of different temperature settings across various clinical tasks. We conducted a retrospective cohort study using electronic health records from the Mount Sinai Health System, collecting a random sample of 1283 patients from January to December 2023. Three LLMs (GPT-4, GPT-3.5, and Llama-3-70b) were tested at five temperature settings (0.2, 0.4, 0.6, 0.8, 1.0) for their ability to predict in-hospital mortality (binary classification), length of stay (regression), and the accuracy of medical coding (clinical reasoning). For mortality prediction, all models’ accuracies were generally stable across different temperatures. Llama-3 showed the highest accuracy, around 90%, followed by GPT-4 (80-83%) and GPT-3.5 (74-76%). Regression analysis for predicting the length of stay showed that all models performed consistently across different temperatures. In the medical coding task, performance was also stable across temperatures, with GPT-4 achieving the highest accuracy at 17% for complete code accuracy. Our study demonstrates that LLMs maintain consistent accuracy across different temperature settings for varied clinical tasks, challenging the assumption that lower temperatures are necessary for clinical reasoning.

## 1. Introduction

Recent advancements in Large Language Models (LLMs) present new opportunities in data analysis, especially in the healthcare sector, where their capability to understand and analyze detailed clinical scenarios offers significant potential [1][2][3][4].

Despite their promise, LLMs are complicated models that pose issues for integration in healthcare workflows, often requiring a deep understanding of the nuances of any decision support tools. Additionally, like other machine learning models, LLMs have certain hyperparameters which can greatly affect their performance. While these hyperparameters have been explored for more general use, there remain open questions about how they affect performance in specialized domains, including healthcare [5][6].

One unique hyperparameter of LLMs is temperature, which is a factor that affects the randomness and originality of the LLM’s output [7][8]. Lower temperature settings are associated with more prototypical and standard outputs, while higher temperature settings are associated with more creative and less predictable responses. Preferences for different temperature settings may be intuitive for certain use cases. For instance, for creative writing, one might prioritize higher temperature settings. For healthcare, however, it is not necessarily straightforward which setting will be most effective, and it may be that different clinical tasks for LLMs may require different settings.

One study [9] has started exploring how temperature influences LLMs in academic scenarios, but research in clinical settings is limited. Our investigation addresses this gap in healthcare by evaluating LLM performance in a range of clinical tasks using real-world patient data, extending from binary classification to regression, to the complex, unstructured task of medical coding. Furthermore, we evaluate the influence of temperature settings on the clinical reasoning capabilities of various LLMs when tasked with interpreting clinical data. This investigation serves to deepen our understanding of LLM functionality in healthcare environments [10][11][12].

## 2. Methods

### 2.1 Study Design

This study employs a retrospective cohort design to investigate the effects of the hyperparameter temperature on the clinical reasoning capabilities of Large Language Models (LLMs). Specifically, the study explores how these settings influence LLM performance across different clinical tasks, including binary classification, regression, and medical coding within a healthcare environment.

We conducted the study using electronic data retrieved from the Mount Sinai Health System electronic health records (EHR). Data was retrieved from Emergency Departments (EDs) of five MSHS hospitals Mount Sinai Beth Israel, Mount Sinai Brooklyn, Mount Sinai Queens Hospital, Mount Sinai West, and The Mount Sinai Hospital, and included ED visits from Jan 2023 to Dec 2023.

Data privacy was ensured through anonymization and compliance with MSHS institutional guidelines according to IRB Protocol (Ethic Committee Name: Mount Sinai Institutional Review Board Approval Code: STUDY-18-00573, Approval Date: June 6th,2021), safeguarding patient information throughout the study.

### 2.2 Cohort Creation

The cohort for this study was constructed using the following inclusion criteria:

Patients over 18 years of age were initially presented in the ED and subsequently admitted to the hospital. Selection of the first physician-authored progress note and the first nurse-authored ED triage note for each patient visit provided the notes were not null and contained more than 20 words. We excluded cases of non-adult patients and patients with missing physician or nurse notes.

### 2.3 Data Collection

The data collected for analysis encompassed both structured and unstructured formats. The structured data included variables such as Sex, Race, Ethnicity, Age, and Discharge Disposition. The unstructured data consisted of clinical notes authored by physicians and nurses. Patient encounters were organized by Arrival-Instant timestamp, serving as the benchmark for initial patient assessments.

Initial Data collection followed systematic steps and was implemented to ensure the inclusion of relevant patient records. Initially, data was retrieved from the database within a specified date range for the year 2023. An age filter was then applied to include only patients who were above 18 years old. The next step involved selecting patients who were admitted through an Emergency Department (ED) visit. Following this, a filter was applied to include only those patients who were hospitalized after their ED visit.

Initial ED diagnoses made by healthcare professionals such as physicians, residents, fellows, and registered nurses were included. Data extraction focused on specific types of ED notes, namely ED Progress Notes and ED Triage/Intake notes and considered it as a filtered dataset.

We sampled a random selection of ED visits from the included hospitals, targeting 250 records of patients per hospital. Given the systematic application of multiple filters, such as age and hospitalization status post-ED visit, the final dataset varied slightly, totaling 1,283 records, well balanced across hospitals.

### 2.4 Outcomes Measures

The study focused on three primary outcomes that were different in their analysis type:

- The first outcome was to predict in-hospital mortality, a binary classification task (1 – Mortality, 0 – No mortality).
- The next outcome was to predict length of stay (LOS), as a continuous numeric prediction in days from arrival to discharge.
- The last task was to determine medical coding accuracy, via the assignment and verification of ICD-10-CM ED primary diagnosis codes based on the clinical narratives and primary diagnosis data.

### 2.5 Model Parameters and Implementation

LLM performance was evaluated under various temperature settings (0.2, 0.4, 0.6, 0.8, 1.0) to assess the impact on the randomness and originality of the outputs. For further configurations such as Top K settings, we employed the API defaults and kept them constant across the different experiments.

**Table 1** provides a detailed overview of the temperature settings and other parameters, ensuring a comprehensive understanding of the configurations used during the experiments.

**Table 1:** Parameters for Text Generation Process.

| Parameter Name | Parameters Value |
| --- | --- |
| Temperature | 0.2, 0.4, 0.6, 0.8, 1.0 |
| Top P (Nucleus Sampling) | 1.0 |
| Max Token Size | 800 Tokens |
| Stop | None |

The study utilized three LLMs: OpenAI’s GPT-4 GPT-3.5 and Meta’s Llama-3-70b for the different experiments as mentioned in **Table 2**.

**Table 2:**
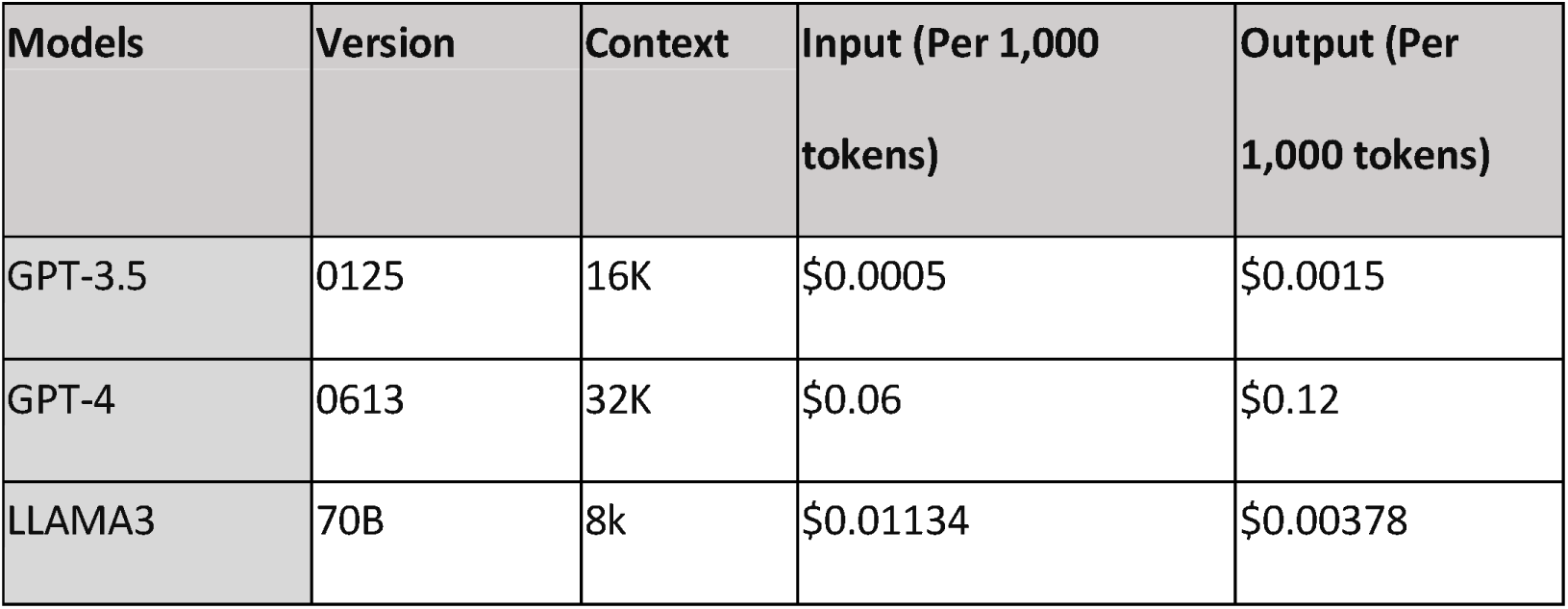
Comparison of Text Generation Models: Context, Input, and Output Costs.

All code and analyses were conducted using Python ver. 3.9. All models were run on the MSHS HIPAA-compliant private Azure cloud instance and accessed via API calls.

Within this infrastructure, we created an Azure AI Studio service under a virtual private network (VPC) to conduct the experiment.

### 2.6 Prompt Designs

The models were prompted to act as clinical practitioners, integrating both structured and narrative data inputs to perform and score predictions on mortality, LOS, and primary diagnosis using a structured JSON format.

The prompt used was:

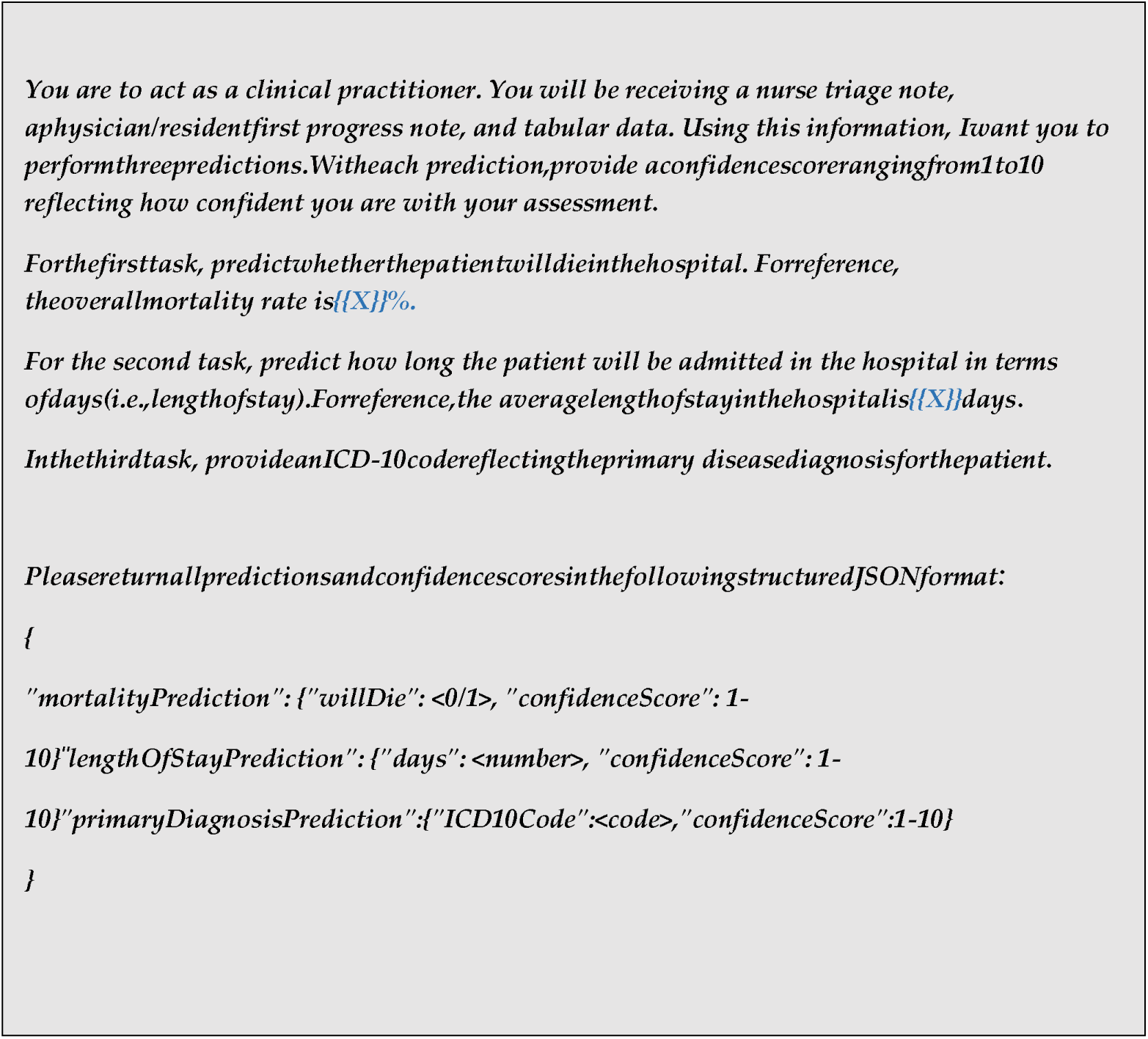

### 2.8 Statistical Analysis

#### 2.8.1 Classification Metrics

Metrics included AUC for probabilities and accuracies for absolute predictions. We have also calculated cut-off-based assessments of sensitivity, precision, specificity, NPV, and F1 score.

#### 2.8.2 Regression Metrics

Mean square error (MSE) and root mean square error (RMSE), which are commonly used metrics for regression analysis, were used to gauge the accuracy of LOS predictions.

#### 2.8.3 ICD Coding Metrics

Accuracy evaluations for ICD coding were segmented into absolute accuracy, and accuracy within the first 2, 3, and 4 digits of the ICD-10-CM codes. This was done to evaluate accuracy with increasing levels of diagnostic specificity (from 2 digits to full code comparison). Confidence scores were quantified by mean, median, and standard deviation measures to evaluate the predictions’ reliability.

## 3. Results

The study focused on predictions of mortality, length of stay, and primary ED ICD-10-CM coding across temperatures ranging from 0.2 to 1.0 for 3 LLMs. These predictions were compared against the clinical ground truths to assess accuracy and confidence across different model settings. The characteristics of the cohort are presented in **Table 3**.

**Table 3:** Cohort characteristics.

| Category | Description | Value |
| --- | --- | --- |
| **Word Count Statistics** | Total Notes | 1,283 |
|  | Average Notes per Patient | 2 Notes |
|  | Mean Words per Note | 101.49 words |
|  | Standard Deviation of Words per Note | 84.38 words |
|  | Minimum Words in a Note | 41 words |
|  | 25th Percentile Words | 52 words |
|  | Median Words | 83 words |
|  | 75th Percentile Words | 118 words |
|  | Maximum Words in a Note | 1,395 words |
| **Distribution of Note Sizes** | Small (less than 52 words) | 317 entries |
|  | Medium (between 52 and 118 words) | 647 entries |
|  | Large (more than 118 words) | 319 entries |
| **Mortality Statistics** | Total Notes or Patients? | 1,283 |
|  | Deceased | 62 |
|  | In-hospital Mortality Rate | 4.90% |
| **Length of Stay** | Mean Length of Stay | 6.93 days |
|  | Median Length of Stay | 4.0 days |
|  | Standard Deviation of Length of Stay | 10.65 days |
|  | Interquartile Range of Length of Stay | 6.0 days |
| **Estimated Tokens** | Total Estimated Number of Tokens | 270,385.25 |
| **Gender Distribution** | Male | 50.43% |
|  | Female | 49.57% |
| **Race Distribution** | White | 39.64% |
|  | Black or African American | 29.83% |
|  | Others | 30.53% |
| **Age Statistics** | Average Age | 64.75 years |
|  | Age Distribution Peaks | 77 years (44), 64 years (43) |
| **Average Age by Race** | White | 67.18 years |
|  | Black | 62.57 years |
|  | Others | 75.55 years |
| **Average Age by Sex** | Female | 66.55 years |
|  | Male | 62.98 years |
| **Age Statistics by Race**<br>**(Extended)** | **Others** | 75.55 years |
|  | Standard Deviation | 15.32 |
|  | Interquartile Range | 22 |
|  | **Black** | 62.57 years |
|  | Standard Deviation | 18.25 |
|  | Interquartile Range | 26 |
|  | **White** | 67.18 years |
|  | Standard Deviation | 19.14 |
|  | Interquartile Range | 28 |
| **Age Statistics by Sex**<br>**(Extended)** | **Female** | 66.55 years |
|  | Standard Deviation | 18.45 |
|  | Interquartile Range | 27 |
|  | **Male** | 62.98 years |
|  | Standard Deviation | 20.11 |
|  | Interquartile Range | 29 |

### 3.1 Classification Task for Mortality Prediction

The evaluation of GPT-4, GPT-3.5, and Llama-3 binary predictions (mortality in hospital yes/no) across varying temperatures revealed consistent accuracy metrics.

Llama-3 persistently scored higher than its counterparts, with accuracies around 90%. In comparison, GPT-4 showed accuracies around 80-83%, and GPT-3.5 showed the lowest accuracies around 74-76% (**Table 4**, **Figure 2**).

**Figure 1:**
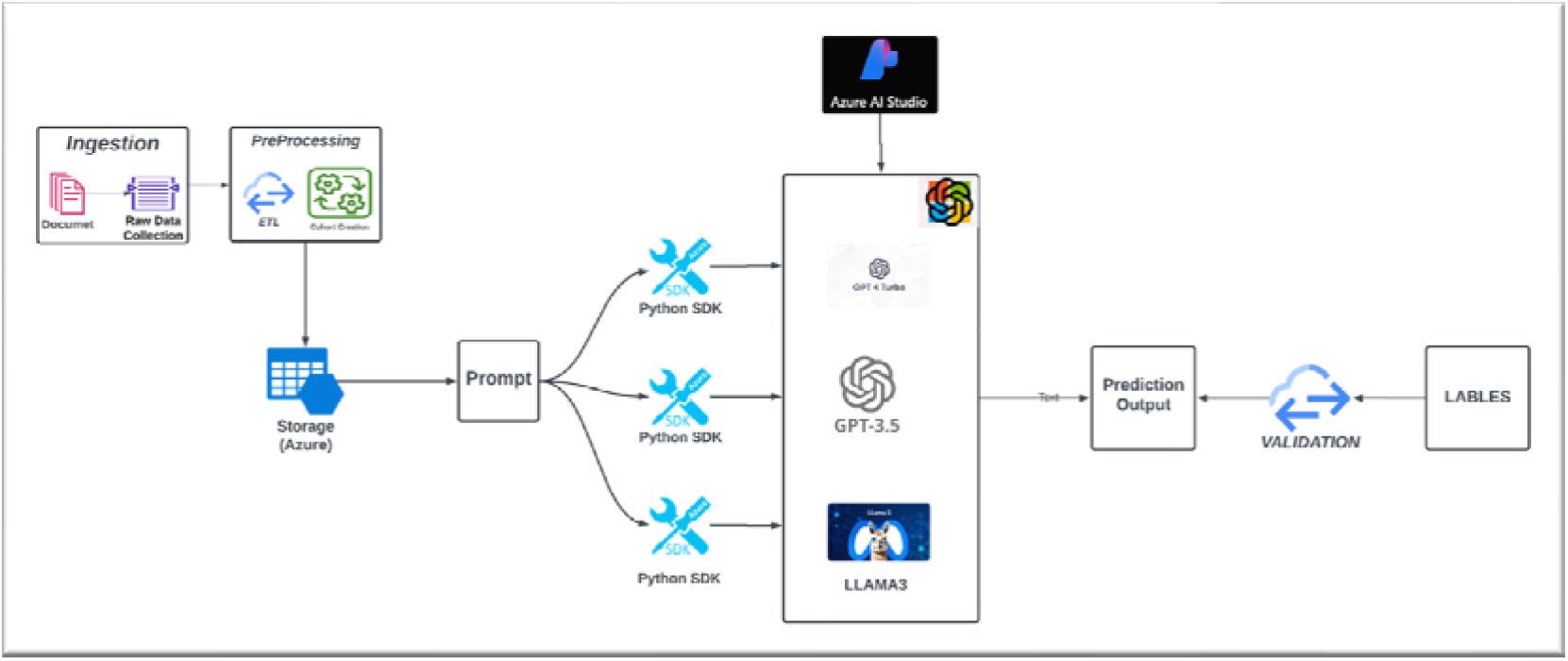
Experiment design with help of Azure AI studio

**Figure 2:**
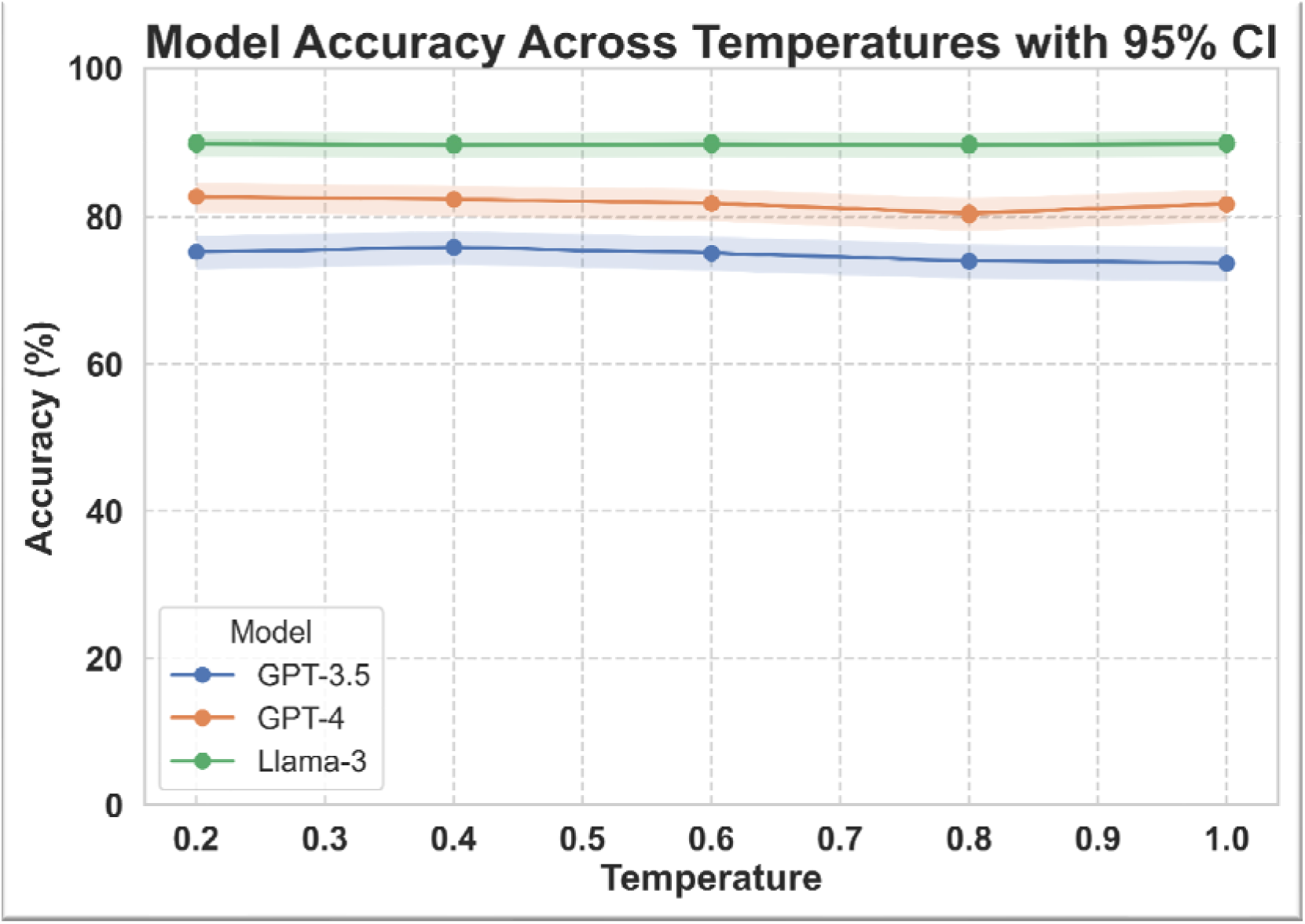
Models’ predicted accuracies stratified by temperatures Abbreviations: CI – confidence interval

**Table 4:** Metrics of LLM models for predicting mortality in hospitals using a binary prediction (0/1).

| Model | Temperature | Accuracy | Precision | Recall | F1 Score |
| --- | --- | --- | --- | --- | --- |
| GPT-4 | 0.2 | 0.83 | 0.27 | 0.48 | 0.35 |
|  | 0.4 | 0.82 | 0.25 | 0.48 | 0.33 |
|  | 0.6 | 0.82 | 0.27 | 0.52 | 0.35 |
|  | 0.8 | 0.80 | 0.26 | 0.55 | 0.35 |
|  | 1 | 0.82 | 0.27 | 0.55 | 0.37 |
| GPT-3.5 | 0.2 | 0.75 | 0.20 | 0.52 | 0.29 |
|  | 0.4 | 0.76 | 0.19 | 0.53 | 0.28 |
|  | 0.6 | 0.75 | 0.20 | 0.52 | 0.29 |
|  | 0.8 | 0.74 | 0.17 | 0.44 | 0.25 |
|  | 1 | 0.74 | 0.18 | 0.49 | 0.27 |
| Llama-3 | 0.2 | 0.90 | 0.45 | 0.20 | 0.28 |
|  | 0.4 | 0.90 | 0.43 | 0.19 | 0.26 |
|  | 0.6 | 0.90 | 0.43 | 0.18 | 0.25 |
|  | 0.8 | 0.90 | 0.43 | 0.19 | 0.27 |
|  | 1 | 0.90 | 0.44 | 0.19 | 0.27 |

When shifting to probabilities, the AUC values for GPT-4 and Llama-3 ranged from 0.713-0.744 for GPT-4 and 0.744-0.755 for Llama-3 (**Table 5**, **Figure 3**). The AUC values remained quite stable across temperatures, indicating consistent discriminatory capabilities. Conversely, GPT-3.5 displayed a slight decrease in AUC with increasing temperatures, from 0.687 for temperature 0.2 to 0.616 for temperature 1.0.

**Figure 3:**
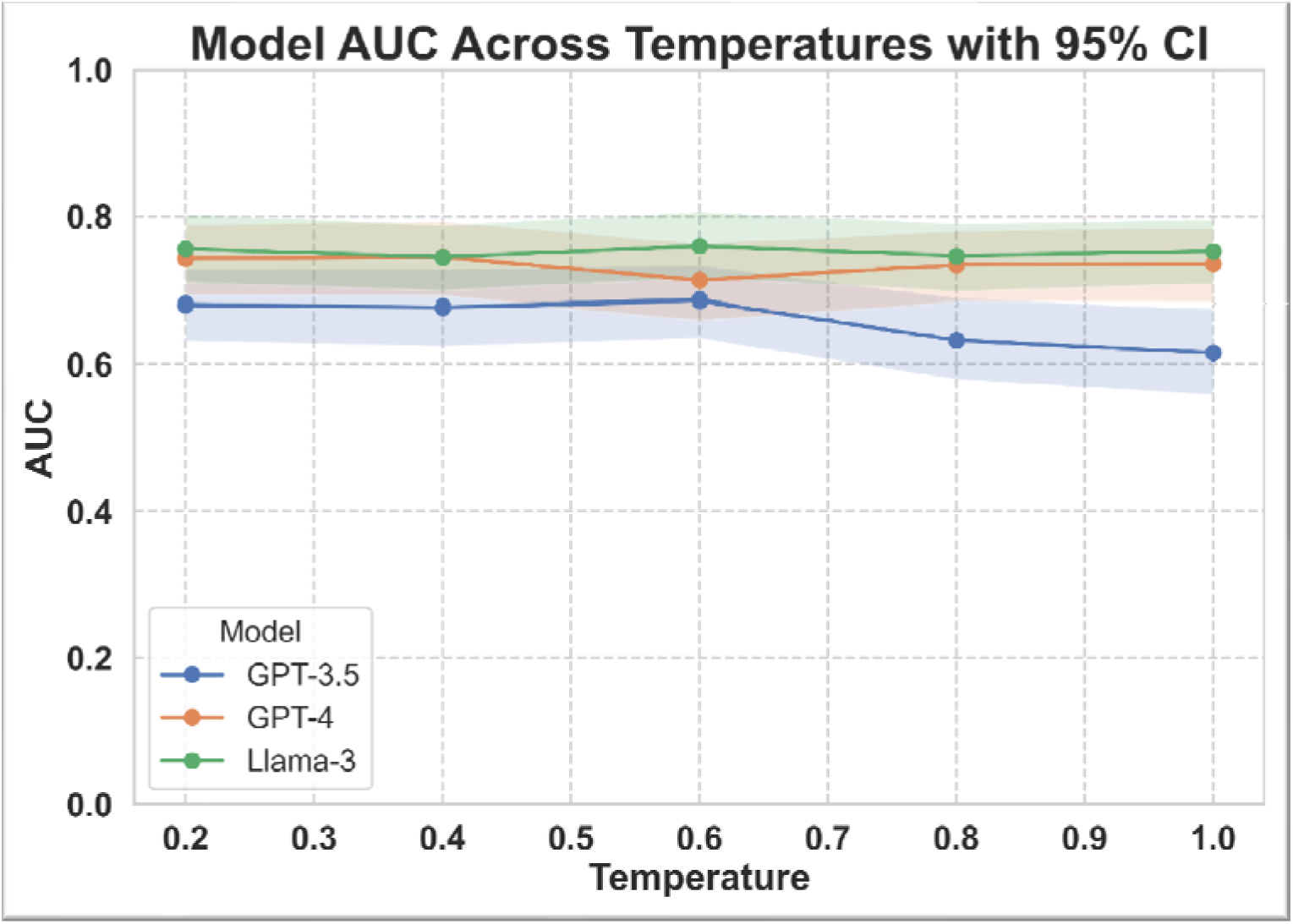
Models’ area under the receiver operating characteristic curve (AUC) stratified by temperatures. Abbreviations: CI – confidence interval

**Table 5:** Metrics Computed Using Youden’s Index. This table displays accuracy, precision, recall, F1 score, and the area under the receiver operating characteristic curve (AUC), derived from the model’s predicted probabilities. Metrics were calculated using the optimal cutoff determined by Youden’s index for each model.

| Model | Temperature | AUC | Accuracy | Precision | Recall | F1 Score |
| --- | --- | --- | --- | --- | --- | --- |
|  |  |  | *For Youden's index probability threshold* |  |  |  |
| **GPT-4** | **0.2** | 0.742 | 0.67 | 0.18 | 0.69 | 0.29 |
|  | **0.4** | 0.744 | 0.66 | 0.18 | 0.75 | 0.29 |
|  | **0.6** | 0.713 | 0.83 | 0.28 | 0.50 | 0.36 |
|  | **0.8** | 0.733 | 0.81 | 0.27 | 0.57 | 0.36 |
|  | **1** | 0.735 | 0.82 | 0.28 | 0.53 | 0.36 |
| **GPT-3.5** | **0.2** | 0.681 | 0.62 | 0.16 | 0.68 | 0.26 |
|  | **0.4** | 0.677 | 0.62 | 0.15 | 0.68 | 0.24 |
|  | **0.6** | 0.686 | 0.55 | 0.15 | 0.77 | 0.25 |
|  | **0.8** | 0.633 | 0.78 | 0.20 | 0.42 | 0.27 |
|  | **1** | 0.616 | 0.78 | 0.20 | 0.40 | 0.26 |
| **Llama-3** | **0.2** | 0.755 | 0.63 | 0.18 | 0.80 | 0.29 |
|  | **0.4** | 0.744 | 0.63 | 0.18 | 0.79 | 0.29 |
|  | **0.6** | 0.759 | 0.66 | 0.19 | 0.81 | 0.31 |
|  | **0.8** | 0.745 | 0.65 | 0.18 | 0.75 | 0.29 |
|  | **1** | 0.752 | 0.64 | 0.19 | 0.79 | 0.30 |

### 3.2 Regression Task – Prediction of Length of Stay in days

The MSE and RMSE metrics computed for the models across various temperature settings show consistency in the model’s performance in predicting the length of stay (**Table 6**). Some slight variations are seen across different temperatures, but no clear trend is demonstrated. Notably, GPT-3.5 generally exhibited slightly higher MSE and RMSE values at most temperature settings, while Llama-3 displayed the lowest RMSE at a temperature of 1.0, suggesting a slight edge in predictive performance at this setting. GPT-4’s results were broadly comparable to those of GPT-3.5.

**Table 6:**
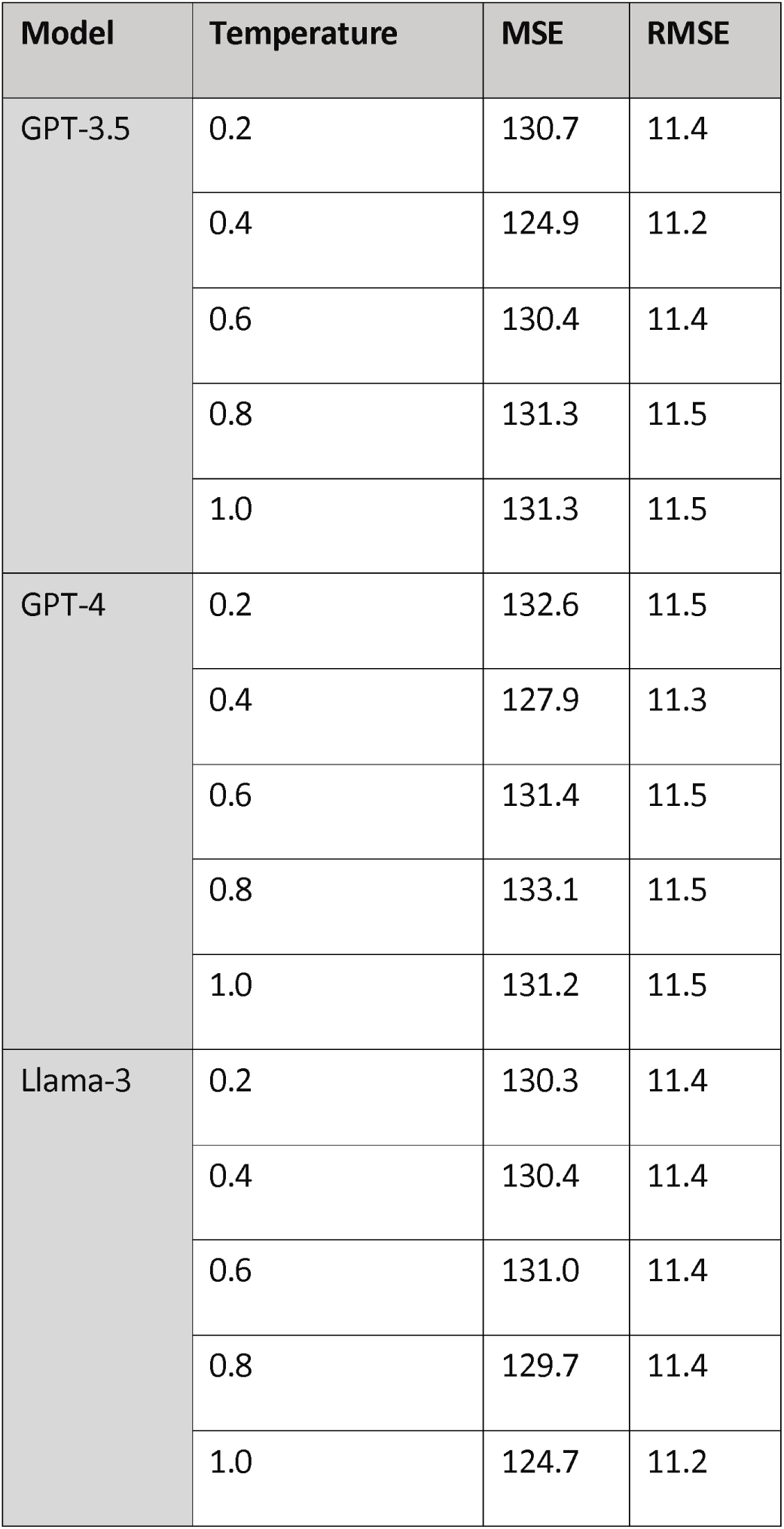
Regression Analysis of Model Predictions for Length of Stay.

The analysis of average confidence scores for LOS predictions made by GPT-3.5, GPT-4, and Llama-3 across varying temperatures indicates a consistent level of confidence across all models and settings.

As depicted in **Figure 4**, confidence scores for each model do not show significant variations with changes in temperature. Specifically, GPT-3.5 and Llama-3 maintain a very close range of confidence across the temperature spectrum, while GPT-4 exhibits similarly steady, albeit slightly lower, confidence levels. This uniformity in confidence suggests that the models’ self-assessment of their predictive capabilities remains stable regardless of temperature adjustments.

**Figure 4:**
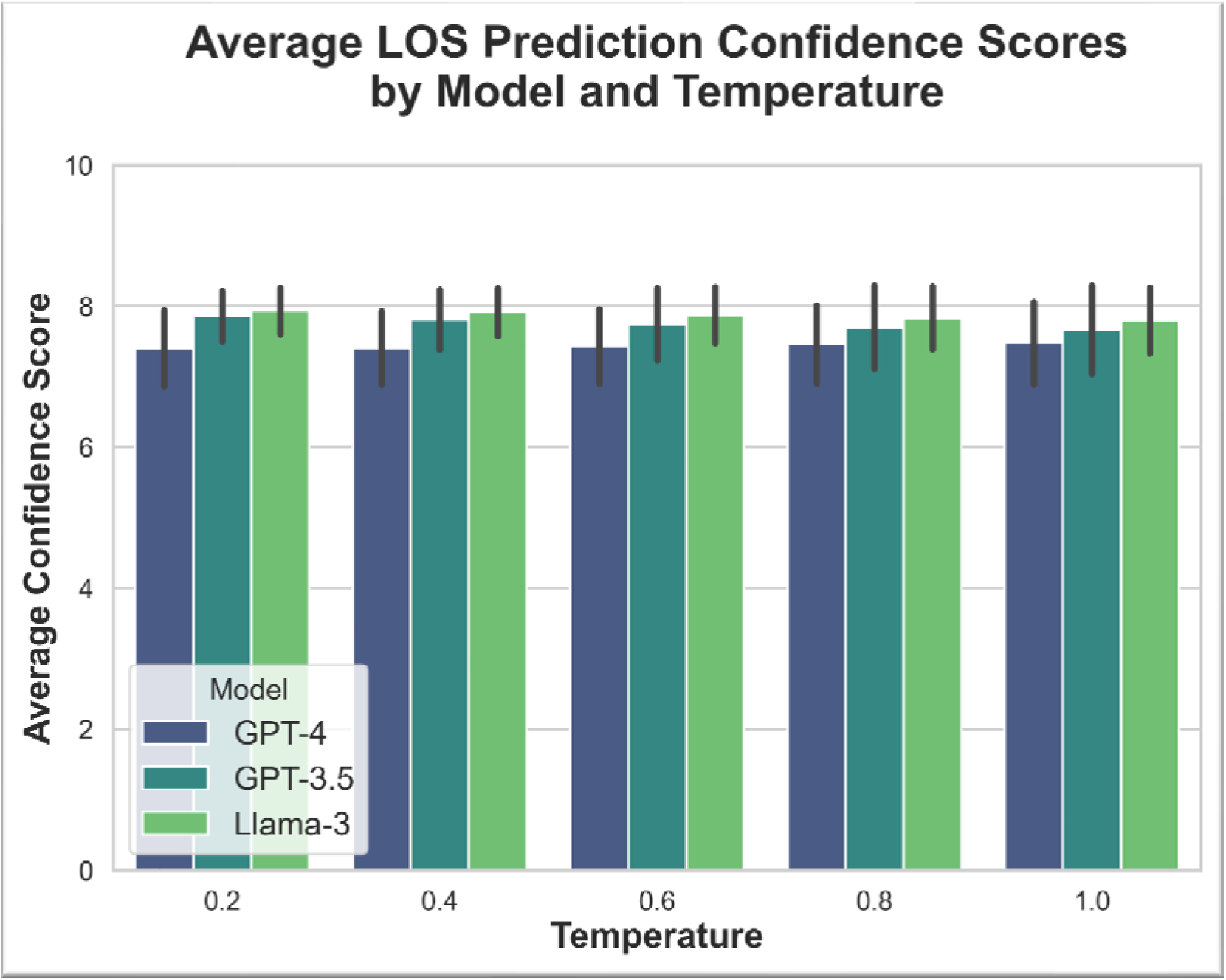
Models’ confidence scores for LOS prediction

### 3.3 Clinical Reasoning - ICD Coding Accuracy

The analysis reveals that all the models exhibit relatively stable performance across different temperature settings (**Table 7**). All the models showed mediocre performance for complete coding accuracy, specifically:

**Table 7:**
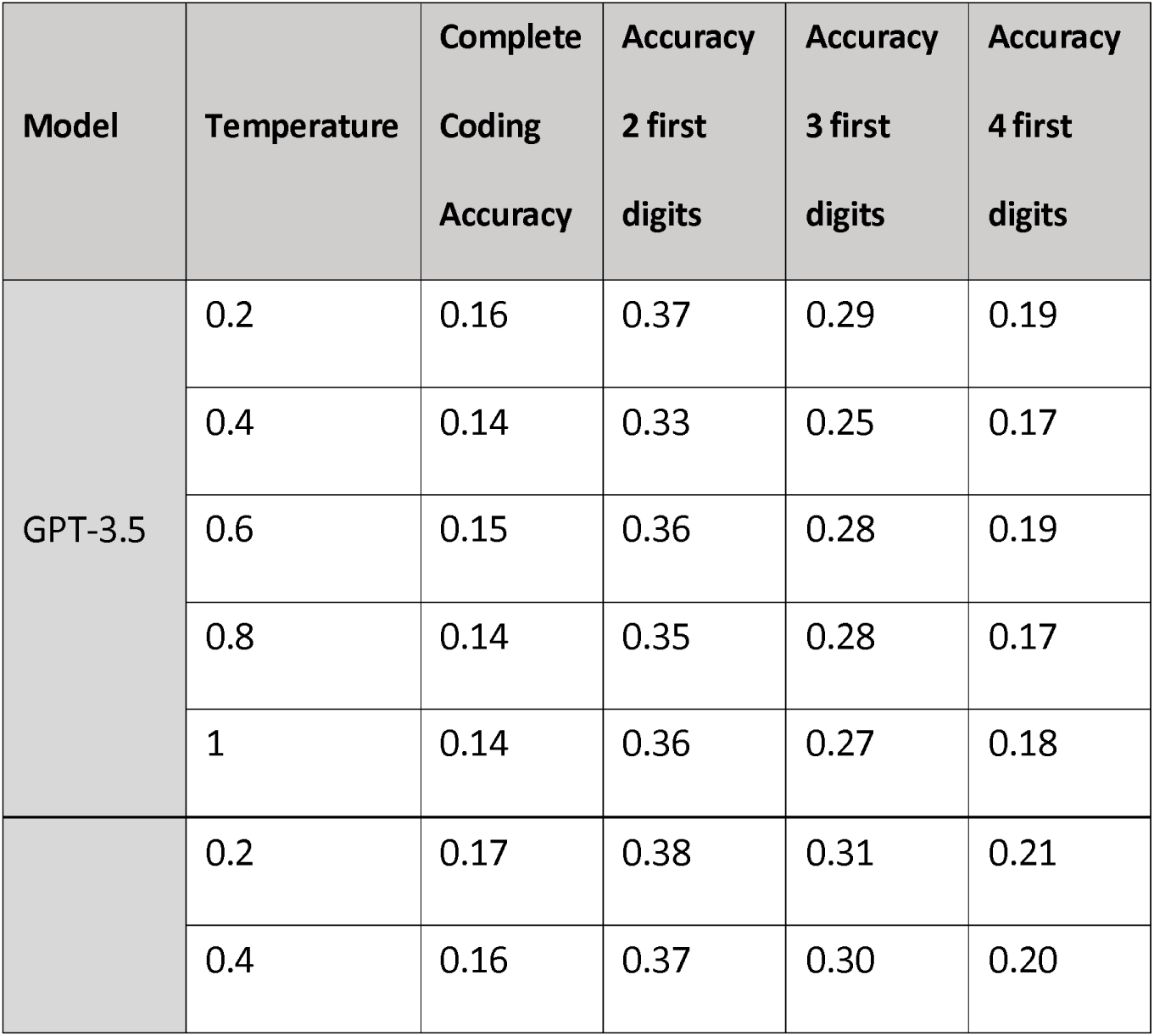
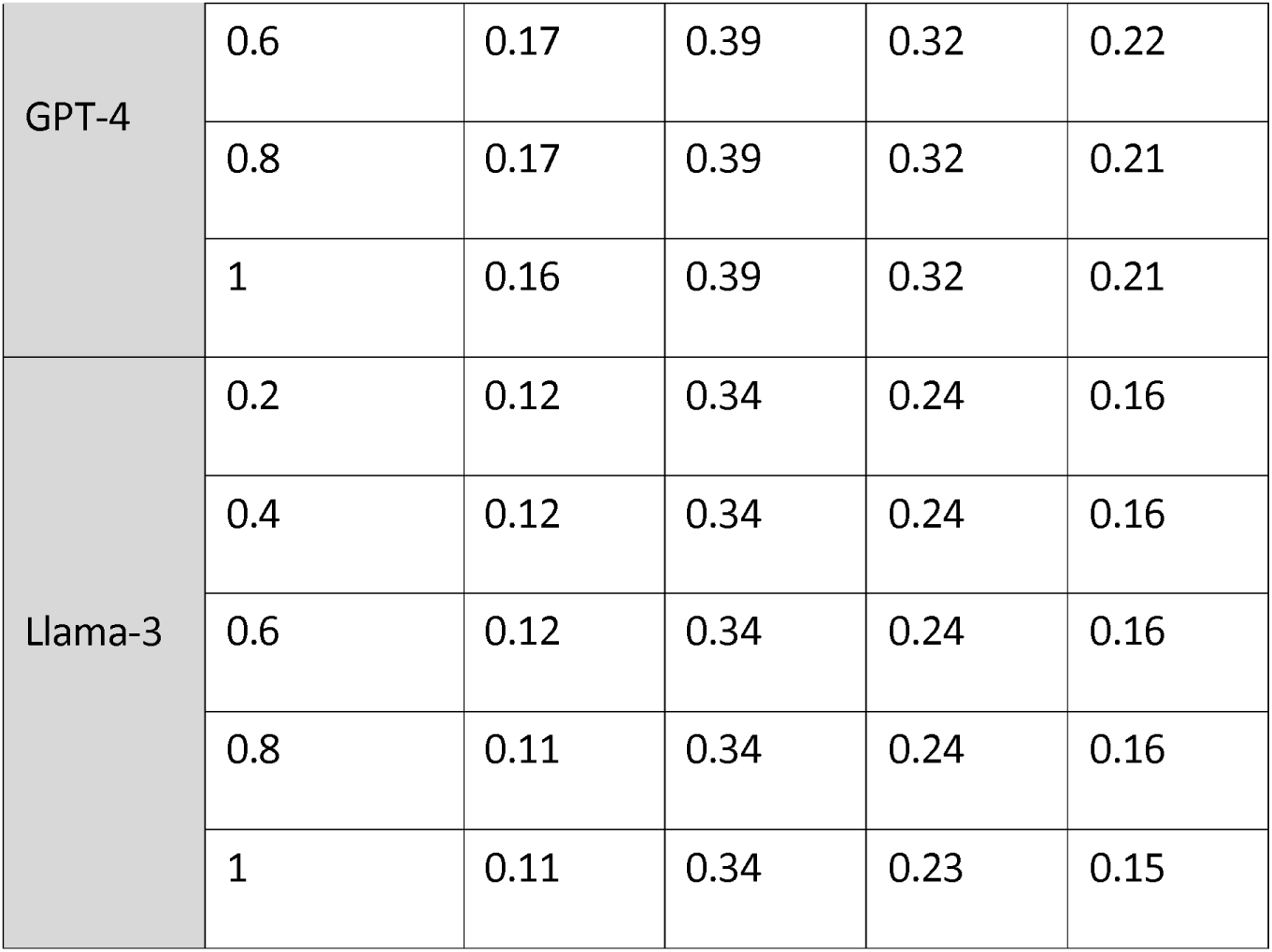
ICD-10-CM coding abilities of the study’s different LLMs across different temperatures.

For *complete code accuracy assessment*, GPT-4 generally demonstrates the highest complete accuracy, peaking at 17%. GPT-3.5 and Llama-3 show lower accuracies, with GPT-3.5 peaking at 16% and Llama-3 reaching only up to 12%. *For the two-digit accuracy assessment,* GPT-4 leads with accuracies of around 39%. GPT-3.5 peaks at 37%, and Llama-3 remains steady at 34%. In terms of the *three-digit accuracy assessment*, GPT-4 peaks at 32%. GPT-3.5’s best is 29%, while Llama-3 averages 24%.

For *the four-Digits assessment,* GPT-4 tops again, exceeding 22%. GPT-3.5 and Llama-3 reach maximums of 19% and 16%, respectively.

Figure 5 illustrates the average confidence scores assigned by each model—GPT-3.5, GPT-4, and Llama-3—for their ICD-10 coding predictions across different temperature settings. Throughout the range of temperatures, each model exhibits relatively stable confidence levels. GPT-4 and GPT-3.5 maintain higher confidence scores compared to Llama-3 across all temperature settings. Notably, confidence scores do not significantly fluctuate with changes in temperature for any model.

**Figure 5:**
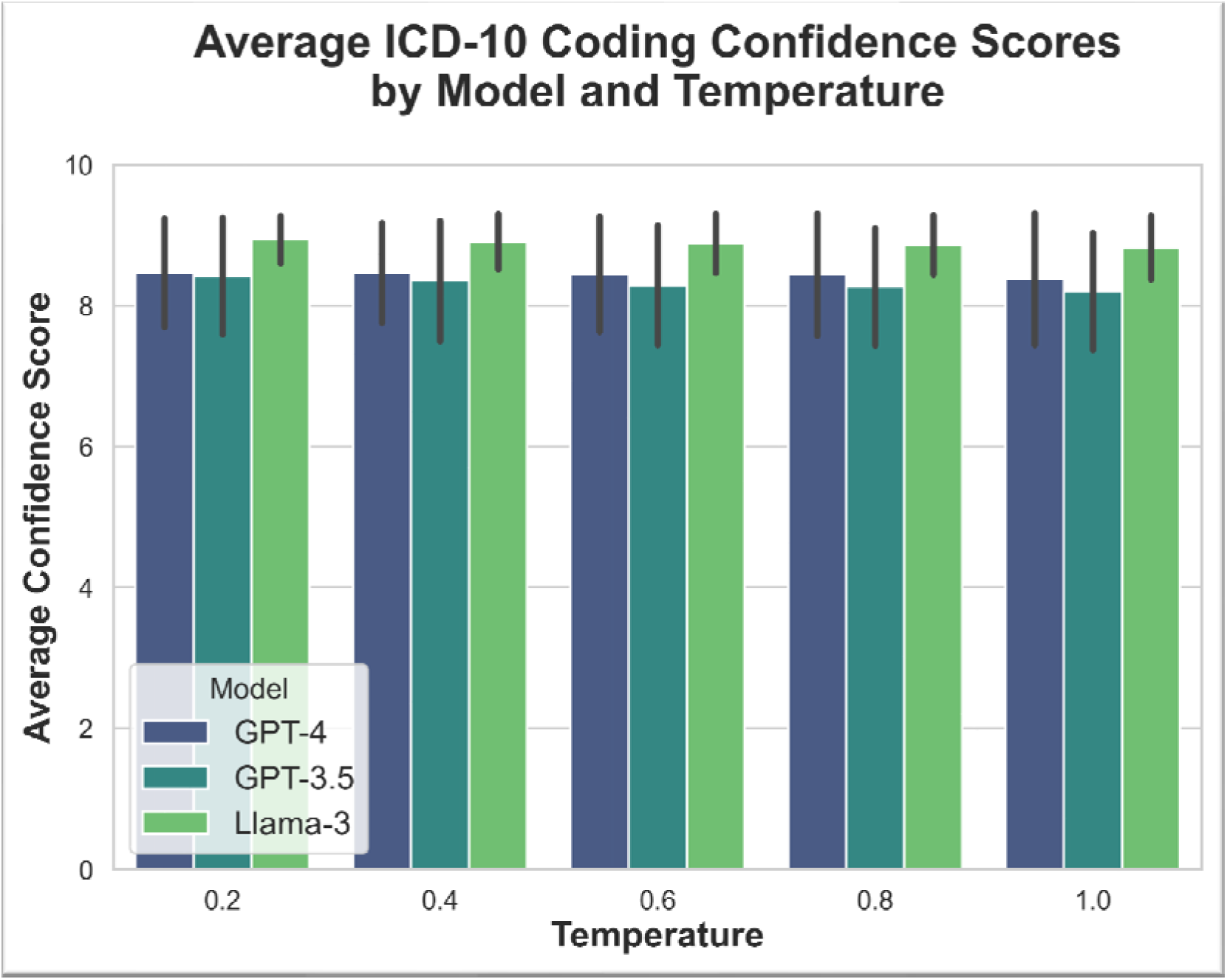
Models’ confidence scores for ICD-10 coding.

## 4. Discussion

Our findings reveal a remarkable consistency in the performance of LLMs like GPT-4 and Llama-3 across a variety of clinical tasks—classification, regression, and the intricate process of ICD coding—regardless of temperature settings. This stability not only underscores the robustness of LLMs in clinical reasoning but also challenges prevailing assumptions about their reliability, suggesting that performance remains solid even as models generate more "creative" outputs at higher temperatures.

This research extends the dialogue initiated by a previous study [9], which explored temperature effects in academic multiple-choice questions contexts, by demonstrating similar stability in the complex realm of real-world clinical data across different clinical tasks.

Another surprising finding of our analysis showed that Llama-3-70b, an open-access model, displayed slightly higher accuracy than GPT-4 in predicting mortality outcomes, highlighting its potential utility in specific clinical tasks. The clinical prediction capacity of LLMs in the ED have been investigated before in few studies [12][13][14][15]. In our previous publication, we have shown GPT-4 had 78% zero-shot accuracy for prediction of hospital admission [12], while Williams et al. have shown GPT-4 had accuracy 89% for classifying acuity level in the ED [13].

However, Llama-3-70b did not perform as well as GPT-4 and GPT-3.5 in the task of ICD coding. This divergence in performance underscores the variability in model effectiveness across different types of clinical data analysis, suggesting that while some models may excel in one area, they may not necessarily perform equally well across all tasks. The low accuracy of LLMs for complete ICD-10-CM coding tasks has been described before [17][18].

Our study underscores the need for developing specific benchmarks in healthcare to assess LLM performance. This entails a focused examination on how LLMs manage unstructured clinical data, which is critical for optimizing their use in healthcare environments. By addressing this gap, the research aims to enhance the precision of LLM applications in patient care settings.

This investigation has limitations. First, it was conducted as a multi-site retrospective study, concentrating on GPT and Llama-3 models and using emergency department (ED) notes as the singular data type. Second, despite covering a range of tasks, many clinical areas remain unexplored. Third, since we evaluated "out-of-the-box" performance, the study did not assess the impact of fine-tuning or Retrieval-Augmented Generation (RAG) on model performance. Also, we’ve limited are research to the usual temperature range of 0.2-1.0. Finally, the number of LLMs available for experimentation is very large. Our study was limited to 3 commonly used ones. These limitations delineate the scope for future studies, particularly in expanding the variety of tasks, and data types, and exploring customization techniques to refine LLM effectiveness.

In conclusion,Our study demonstrates that LLMs maintain consistent accuracy across different temperature settings for varied clinical tasks, challenging the assumption that lower temperatures are necessary for clinical reasoning.

## Data Availability

All data produced in the present study are available upon reasonable request to the authors

